# Dynamic PET imaging predicts broadly neutralizing antibody distribution and HIV prevention efficacy

**DOI:** 10.1101/2025.11.26.25341044

**Authors:** D.H.T. Chu, L. Kuo-Esser, D.R. Beckford-Vera, R.R. Flavell, Y. Seo, H. VanBrocklin, T.J. Henrich, A.N. Deitchman

## Abstract

HIV continues to impose a significant global burden despite success of effective antiretroviral therapy and pre-exposure prophylaxis (PrEP), due to stigma, access, and continuity of treatment that impact real-world effectiveness. Broadly neutralizing antibodies (bNAbs) are novel monoclonal antibody therapeutics being studied for long-acting ART and PrEP, and part of curative HIV regimens. Clinical translation for dose optimization remains a major challenge for the development of these therapeutics. Further, methods to characterize tissue levels of these agents are limited and often impractical due to the need for invasive tissue biopsies. Here we demonstrate the ability of serial whole body positron emission tomography (PET) imaging, following microdosing of the bNAb VRC01 in people without HIV, coupled with physiological-based pharmacokinetic (PBPK) modeling, to accurately predict therapeutic plasma, tissue exposure and prevention efficacy in two major VRC01 prevention trials. Based on our PBPK model, we determined a >51-fold anorectal tissue VRC01 level:inhibitory concentration (IC_80_) target would achieve 90% prevention efficacy compared to >200 based on plasma levels in the primary trial analysis. Thus, these PET-PBPK approaches are promising for noninvasive determination of bNAb penetration more closely linked with concentrations needed to prevent virus acquisition, and may be leveraged to improve efficient development of bNAbs.

## Introduction

Human Immunodeficiency Virus (HIV) remains a global public health concern. As of 2023, there are approximately 40 million people living with HIV (PWH), including an estimated 1.3 million new infections (1). With widespread use of highly active antiretroviral (ARV) therapy in PWH, the advent of pre-exposure prophylaxis (PrEP) strategies, and increased use of barrier prevention methods, the incidence of HIV infection has been reduced since 2010 by 39%(2). Stigma, access to therapy, and issues with treatment continuity reduce real-world effectiveness of these approaches, necessitating long-acting prevention and treatment options to reduce new infections and reduce associated morbidity and mortality of PWH(3–6).

Broadly neutralizing antibodies, bNAbs, are long-acting antiviral agents targeted conserved epitopes on the HIV envelope (7–9). Further, unlike other ARVs, bNAbs show promise to potentially reduce the viral reservoir(10), and may be a key component of a future curative HIV regimen. Unlike approved ARVs, which primarily inhibit integration and proliferation of HIV in host cells, bNAbs work through direct targeting of specific epitopes on the HIV envelope, thereby neutralizing the virus and preventing it from fusing with and entering the host’s cells. It has also been hypothesized that bNAbs may potentiate immune mediated anti-HIV responses(11). Thus, ongoing clinical trials investigate its use as long-acting prevention or treatment with favorable safety and pharmacokinetic profiles, as well as a possible component of an HIV cure regimen(8).

There is a need to develop translational frameworks to evaluate potential of bNAbs for clinical use. Their action in tissue to prevent, treat, or cure HIV is critical to inform dosing and large proof of concept trials. Tissue biopsies are invasive and not feasible for many tissues of interest. Further they are required specialized expertise to obtain and usually limited to a single time-point(12). To overcome the limitation, positron emission tomography-magnetic resonance (PET-MR) methods have been developed as a less invasive method that can be used to obtain serial measurements to determine drug pharmacokinetics in tissues of interest(13, 14). Studies on pharmacokinetic modelling leverage PET-based data in humans for studies in lung or brain distribution of drugs for tuberculosis treatment(15, 16). The models could provide insights on tissue level pharmacokinetics to inform bNAbs drug development prior to larger clinical trials.

The Antibody Mediated Protection (AMP) trials, which evaluated the safety and efficacy of the bNAb VRC01, have provided critical insights into the potential role of bNAbs in preventing HIV-1 acquisition(17). While VRC01 did not significantly reduce overall HIV-1 infection rates, it demonstrated high efficacy against specific HIV strains that were sensitive to the antibody, offering proof-of-concept for bNAb-based prophylaxis(18). The AMP trial mainly focused on blood concentrations. Their analysis indicated 200-fold higher concentration than the bNAb inhibitory concentration to suppress 80% of viruses (IC_80_) to give 90% clinical prevention efficacy(19). This discrepancy is likely due to lower tissue penetration at the site of infection (i.e. most often the anorectal tissue) for which approximately 10-fold lower exposure was observed(20). While informative, this data does not clearly delineate impact of changes in bNAb exposure over time in tissue and the relationship most relevant to clinical prevention efficacy.

In this study, to better understand and optimize future bNAbs protective potential, we built a physiologically based pharmacokinetic (PBPK) model from dynamic PET imaging data following microdosed VRC01. We observed that blood and tissue exposure observed could be scaled to predict plasma and tissue pharmacokinetics at therapeutic doses used in the AMP trials. Further the PET ratio and PBPK derived protection efficacy, transformed from the plasma model of protection efficacy, we more closely linked to the expected PK:IC_80_ ratio required to prevent infections. This work establishes PET-based microdose combined with PBPK modeling as a promising framework for development of bNAbs for HIV prevention.

## Results

### Conversion of radiotracer signal uptake to concentration

The available data with measured concentrations of the percent injected drug per mL of blood measured were plotted against the matched mean PET standardized uptake values (SUVmean) in blood (Fig. 1). Fitted linear regression equation *y* = 0.0011*x* + 0.0013 was used to estimate the concentration (in % g VRC01 / g) in each organ from the corresponding SUVmean.

**Figure 1.**
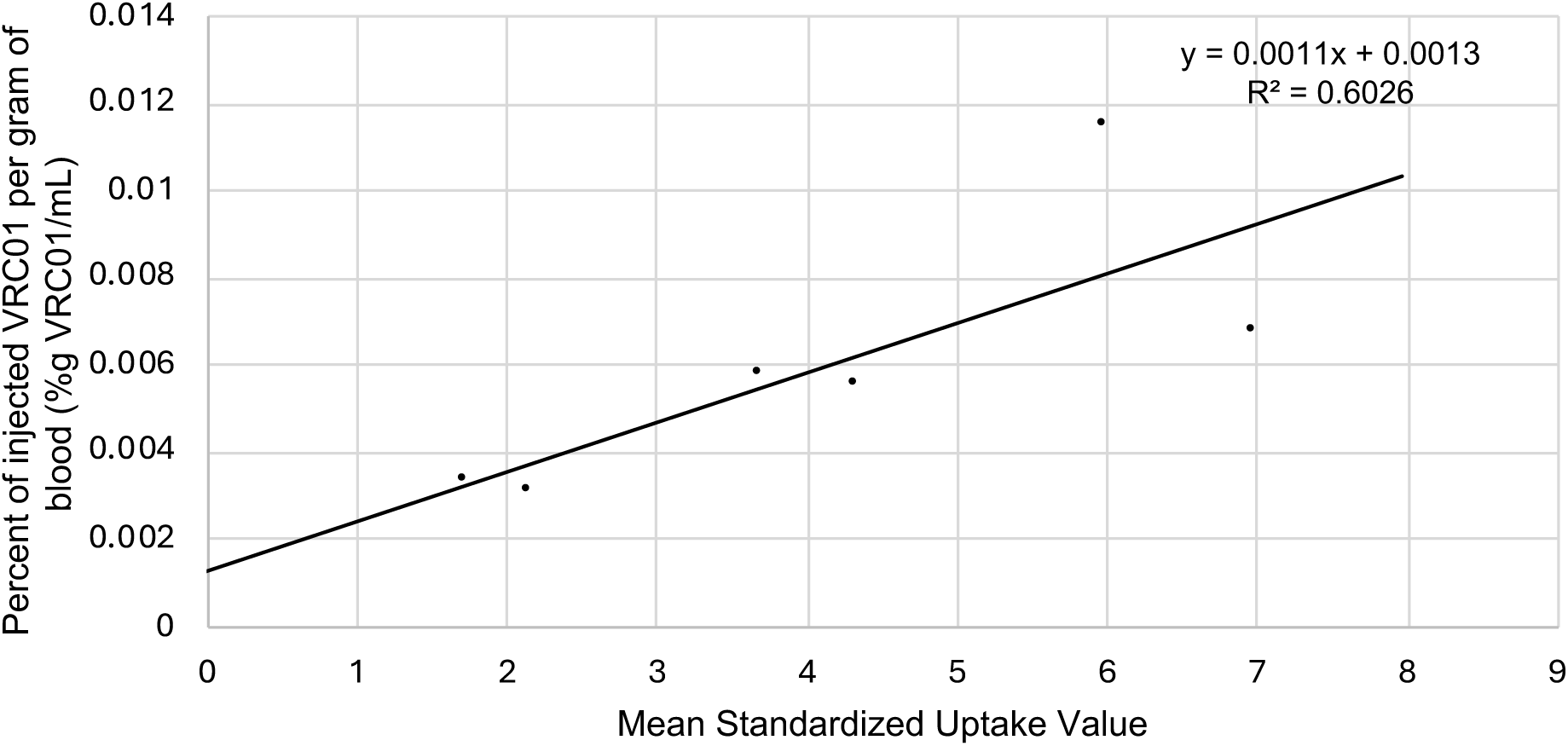
Blood PET uptake linked to measured VRC01 blood level. Correlation plot of mean radiolabel uptake signal in blood versus percent of injected drug per gram of blood. Points are raw data points and solid line represent linear regression.

### PBPK Model for VRC01

A physiological based pharmacokinetic (PBPK) model was constructed from PET-derived VRC01 concentrations in blood, gut/anorectal, liver and spleen (Schematic in Fig. 2). Due to the small sample size of the study, where inter-individual variability (IIV) could not be characterized, and the proportional error used to characterize residual variability. The residual error was described using a proportional error model for all four compartments.

**Figure 2.**
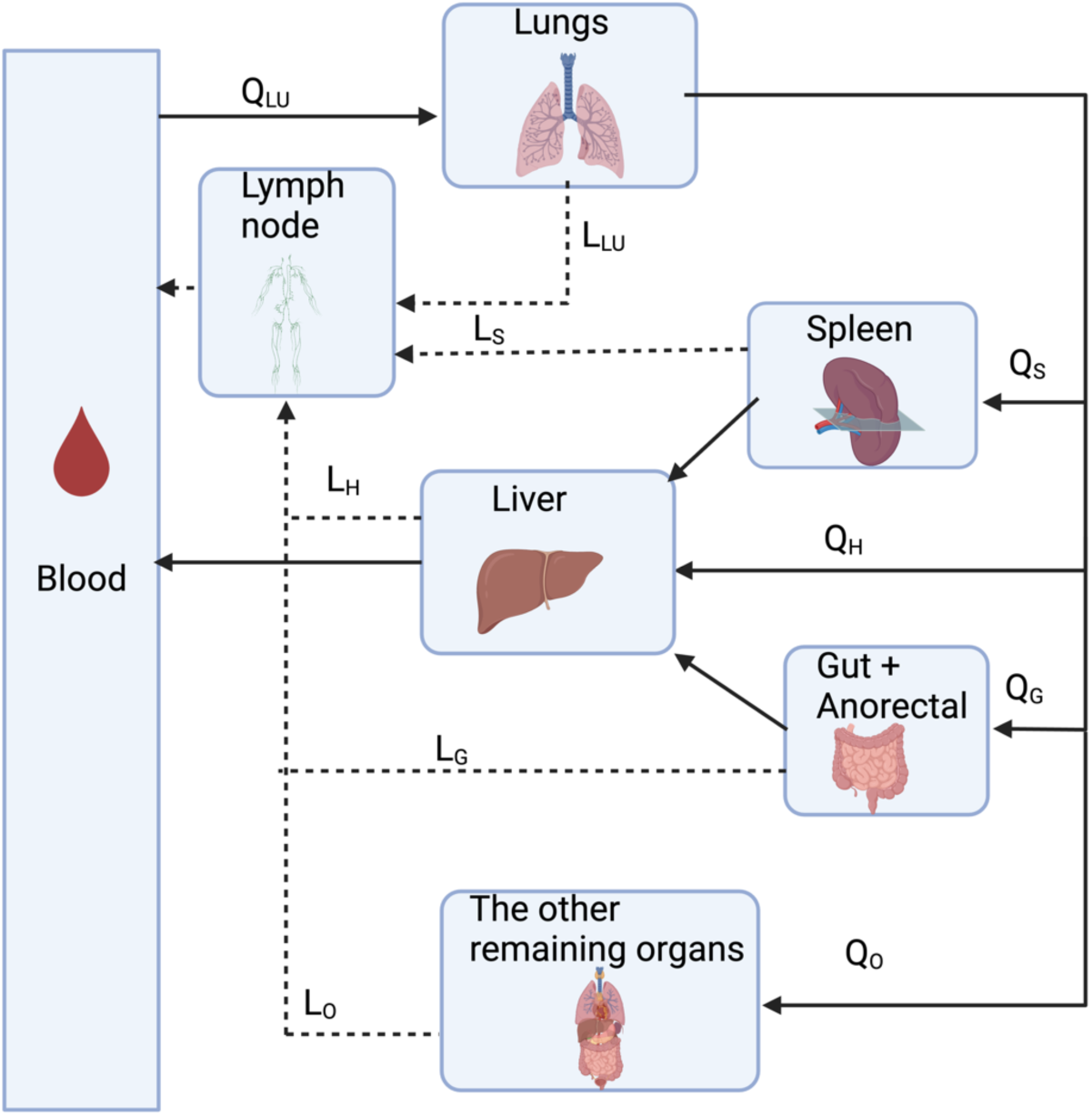
Schematic of PBPK proposed model for PET imaging data. Each compartment represents a physiological organ or space. Abbreviations: Q_Lu_: Blood flow to lungs, L_Lu_: Lymphatic flow from lungs, Q_S_: Blood flow to spleen, L_S_: Lymphatic flow from spleen, Q_H_: Blood flow to liver, L_H_: Lymphatic flow from liver, Q_G_: Blood flow to gut and anorectum, L_G_: Lymphatic flow from gut and anorectum, Q_O_: Blood flow to other remaining organs, L_O_: Lymphatic flow from other remaining organs

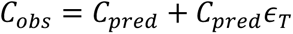

C_obs_ and C_pred_ represent the observed concentration and the predicted concentration, and ϵ_T_ represent the proportional random error of the tissue. The estimated parameters are shown in Table 2, and the visual predictive check between the simulated data and actual data is shown in Fig. 3.

**Figure 3.**
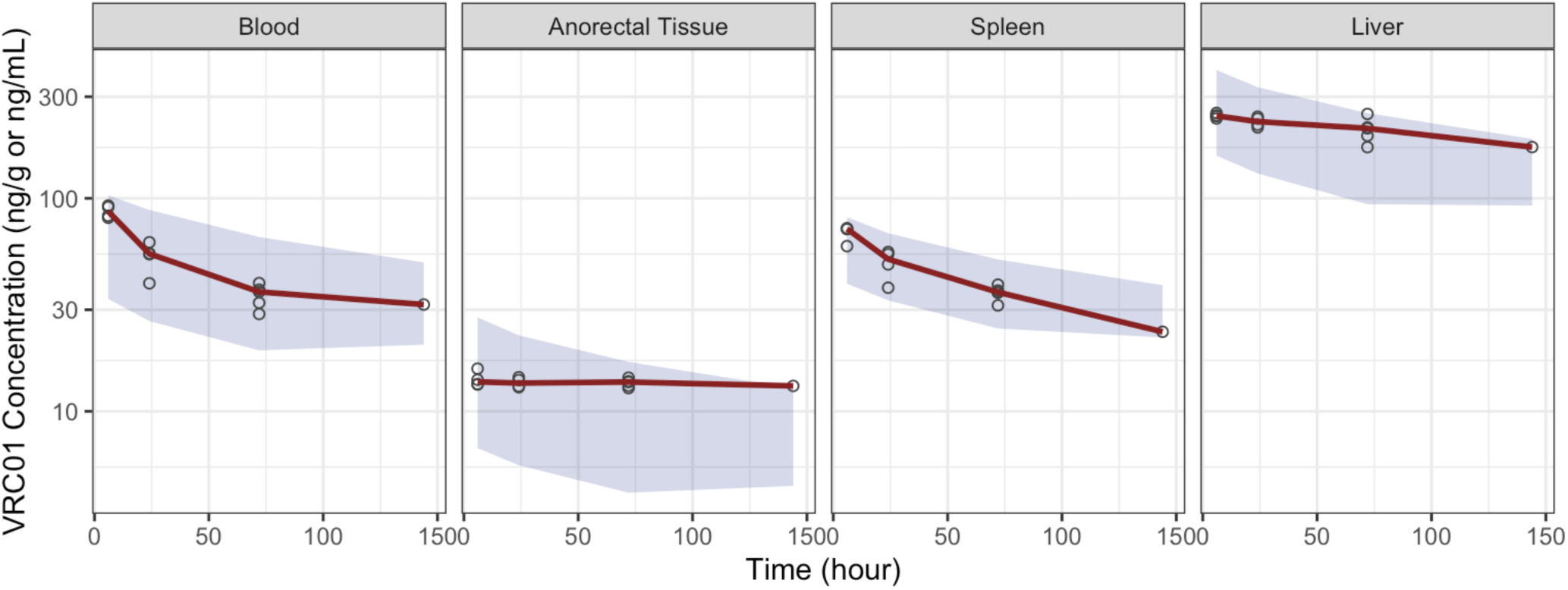
PBPK model predicts PET-based blood and tissue VRC01 pharmacokinetics. Observed VRC01 concentrations (points and mean line) with 90% prediction interval based on 1000 simulations. Each panel represents a different matrix or tissue.

### PET based PBPK model following microdosed VRC01 predicts clinical dose pharmacokinetics in plasma and tissue

Simulated blood PK from our PET based PK model consistently predicted observed AMP trial plasma PK levels for both 10 and 30 mg/dose levels for 10 infusions (Fig. 4). Predicted tissue levels were approximately 10-fold lower for anorectal tissues(17).

**Figure 4.**
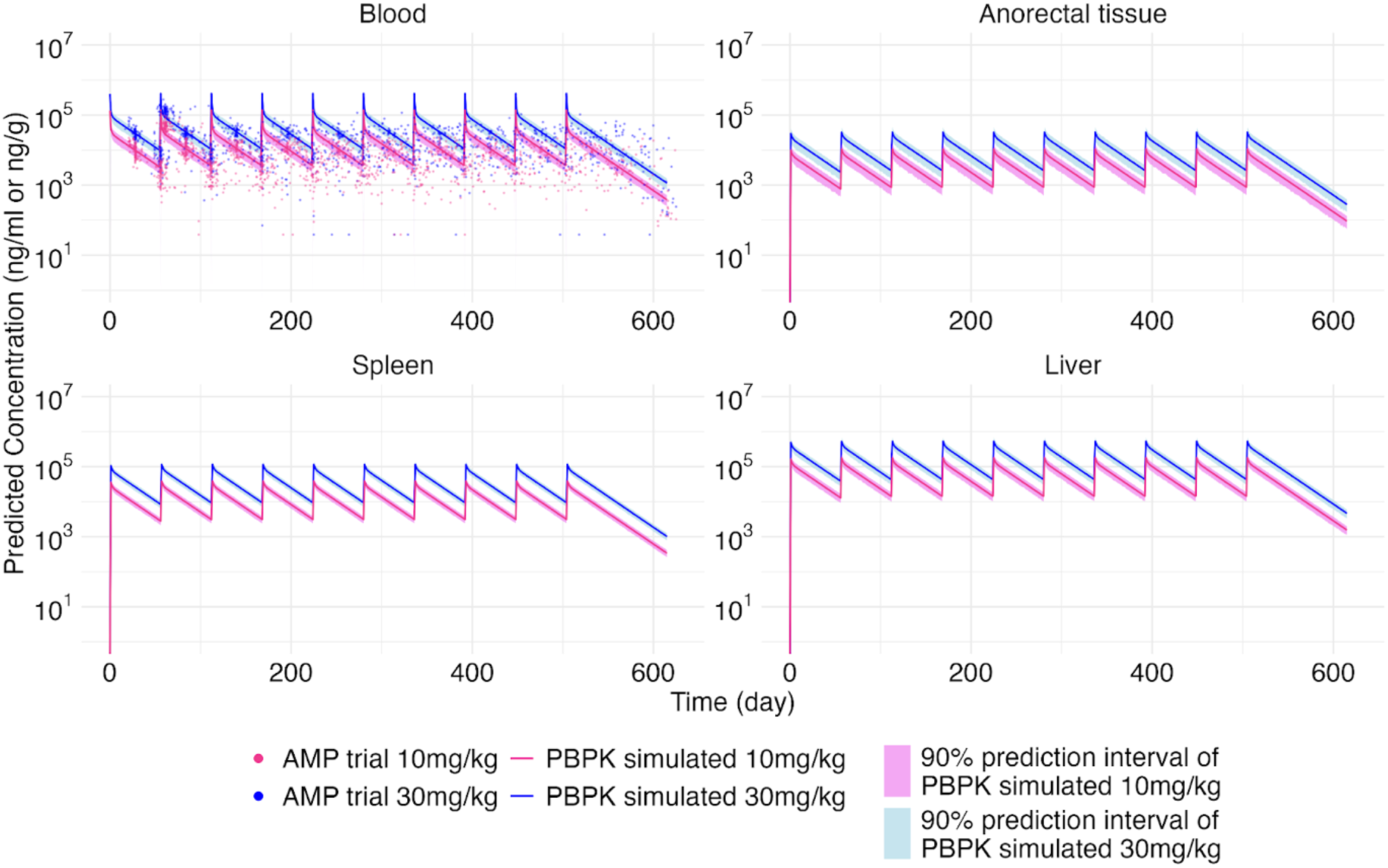
PET based PBPK model predicts AMP VRC01 clinical trial pharmacokinetics. PBPK simulated concentration (ng/mL in blood, ng/g tissue) across blood and tissue (population average prediction [lines], with shaded 90% prediction interval), overlaying with the PK data from AMP VRC01 trials.

### Prevention Efficacy

The modeled gut/anorectal concentration was implemented in the non-human primate model with the daily simulated PK in anorectal tissue divided by the geometric mean of IC_80_ of HIV from participants who became infected with HIV during the AMP trials, compared to using the anorectal:blood ratio of SUVmeans from each tissue PET measurement (Fig. 5).

**Figure 5.**
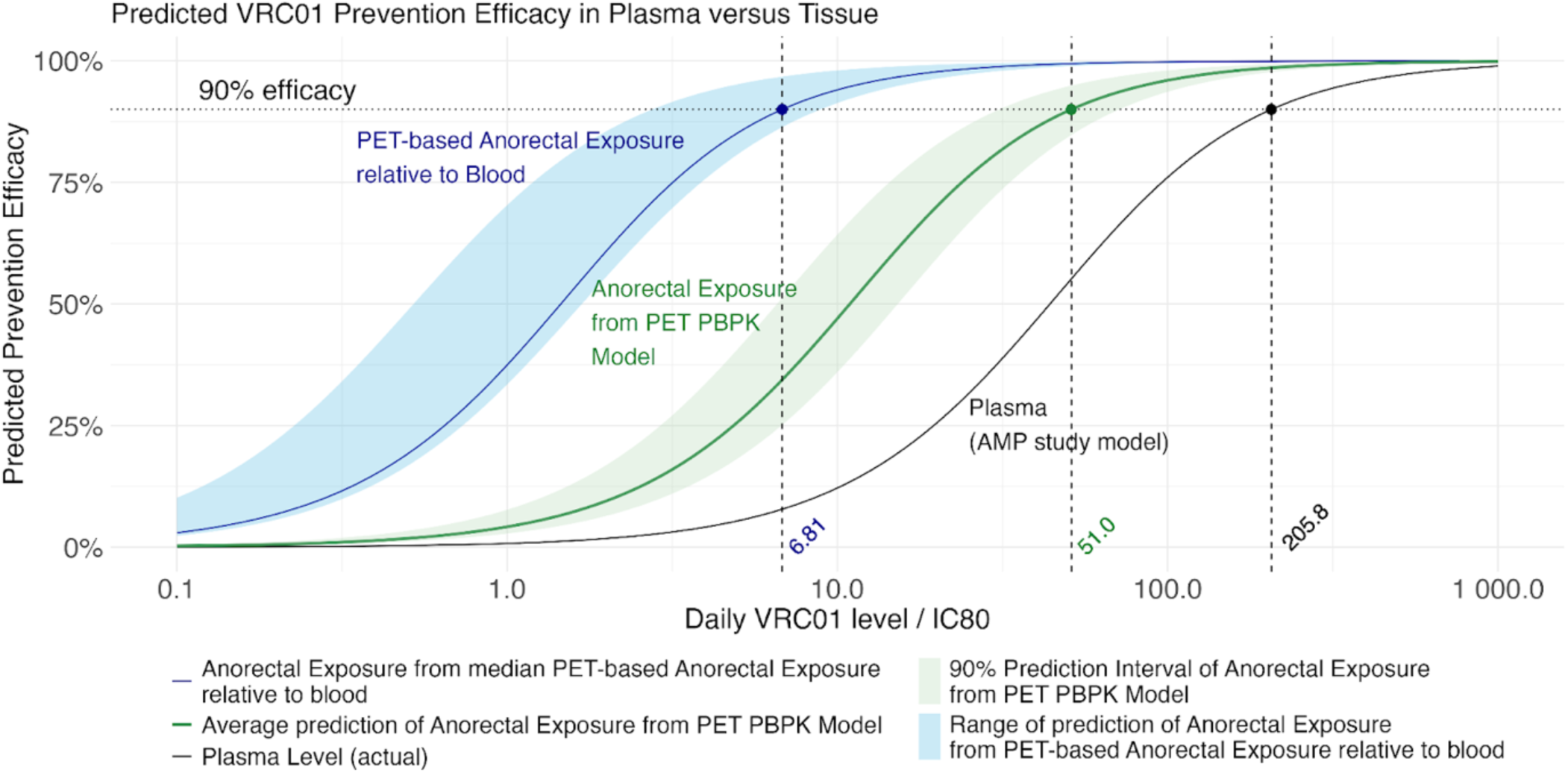
PET-based tissue distribution more closely linked to prevention efficacy. Predicted prevention efficacy as a function of average daily VRC01 trough:viral susceptibility (IC_80_) ratio. Plasma exposure (black line) is based on the model from the AMP VRC01 prevention trials. PET ratio-based anorectal exposure (blue) is predicted from the ratio between anorectal tissue to blood tracer distribution. PET-PBPK anorectal exposure (green) is based on model predicted exposure. The VRC01 trough level:IC_80_ required to achieve a median prevention efficacy of 90% is noted for each method.

The result showed that compared to using plasma concentration, which requires a concentration of 206-fold from IC_80_ to achieve 90% prevention efficacy, the anorectal tissue concentration modelled by the PBPK requires a mean of 51-fold (range: 10.4 - 95.0). Using anorectal tissue:blood ratio of SUVmean from PET (median: 30.2 [range: 24.4 – 79.0]), yields a lower value (median: 6.81-fold [range: 2.61 – 8.43]) demonstrating the need for comprehensive PBPK analyses to characterize differences in distribution kinetics.

## Discussion

This study demonstrates that non-invasive PET imaging with radiolabeled bnAbs may serve as a tool in future drug development to assess bNAb distribution and potential protection efficacy in target tissues(30, 31). Although popPK based on AMP trial data have provided valuable information into the relationship between plasma VRC01 concentrations and HIV prevention efficacy, our study addressed the gap by focusing on the anorectal tissue as a target tissue in preventing HIV(19, 32).

PET based measurements while requiring specific expertise to develop tracers, execute, and interpret studies are likely cost-effective in understanding distribution of candidate therapeutics with targeted action in tissues. Due to cost and radiation exposure, testing is usually limited to a small number of participants, but yet still, as in this study can provide valuable insights for drug development. Even more, this work demonstrates the ability to scale from microdoses of antibody to predict plasma and tissue concentrations at therapeutic, clinical doses.

Despite conversion of SUVmean to VRC01 concentration did not yield a high R^2^ value (0.60), likely due to a lower sample size (n=6), the expected values and resulting PBPK model provide accurate prediction of clinical trial data. It is important to note that the assumption of the same conversion between blood and other organs may be important to investigate in the future when tissue and PET samples are available from the same study. However, given that this is first-in-human data on VRC01 with PET scan, a moderate R^2^ is still informative to serve as a valuable foundation.

We decided to use distribution coefficient, clearance rates, and fraction of lymphatic drainage that contributes to blood flow (FR) as parameters to estimate, reproducing the methodology used by Bae and colleagues (22). These parameters incorporate antibody properties instead of solely physiology (e.g., blood flow rate or lymphatic flow rate in different organs). We assumed the lung distribution coefficient was the same as what was estimated with trastuzumab due to the lack of lung data in our PET imaging studies. Further challenges in differentiation fecal matter from gut tissue in the scan may not represent the tissue concentrations(22). Fortunately, findings of tissue penetration to gut biopsies from the AMP studies with similar ratios to our PET-based findings (∼10:1 plasma to gut VRC01 levels) are encouraging that these values accurately represent gut bNAb levels(33).

When it comes to the estimated parameters, the distribution coefficient and FR showed a relatively precise estimate, compared to clearance, which resulted in a wide 95% confidence interval. This could be explained by the differing elimination rates across different tissues or the need for later timepoints during VRC01 washout(34). The clearance was assumed to be in a non-specific disposition as in individuals without HIV, there is assumed to be no target-mediated elimination(35–37).

Visual predictive checks (VPC) may provide an acceptable fit of the data even with a low sample size. It was interesting to see that the gut/anorectal concentration remained relatively flat throughout the study period. Although the model assumed a linear elimination from the tissue, the mechanism of elimination and distribution within future studies is warranted.

Our approach to simulate a 1000-person virtual population with each individual weighing 70 kg was based on the assumption that our PBPK model did not include weight as a covariate on clearance due to a low sample size. We expect body weight and sex may influence clearance and should be the topic of further studies for this methodology(38).

When we linked the anorectal tissue PK with prevention efficacy model, we used published models developed based in part on data in non-human primates(19, 28). Their finding was that the predicted serum concentration of 200-fold higher than that concentration required to reduce infection by 80% *in vitro* (IC_80_) in order to achieve 90% predicted prevention efficacy. Our approach examined the ratios of the SUVmean in PET studies, and those ratios from PBPK predictions between anorectal tissue and blood to predict the predicted prevention efficacy. The simple ratio method used gave a much lower VRC01 level to IC_80_ required to achieve 90% prevention efficacy, compared to PBPK and the published ratio. As PBPK incorporates physiology that gives a full picture of the bNAb pharmacokinetics in the human body, using a simple ratio from discrete time points may result in overestimating the prevention efficacy with respect to ratio between daily VRC01 level and IC_80_.

Moreover, the concentration from biopsies of rectal tissue, taken from 4 - 13 days after administration, was found to be approximately 10-fold (range of ratio calculated based on IQR: 5.6 - 40.6) less than the serum concentration in males administered with VRC01 at a dose of 10mg/kg and 30mg/kg, after total protein normalization (20). This justified our simulated results of a median-to-median blood-to-anorectal ratio of 4.07 (range of ratio calculated based on IQR: 1.34 - 12.1) in 4 - 13 days after administration, demonstrating the PBPK is potentially less biased compared to a simple ratio method that suggested a median of 30.2 (range: 24.4 to 79.0)-fold difference. A similar result was observed consistently in both VRC01 (16.38% penetration from blood to rectal tissue, IQR: 8.51 - 16.64%), compared to the PBPK model showing 24.72% penetration (IQR: 19.90 - 30.73%) after 5 - 6 weeks(33). One thing to note is that although the IQR for our PBPK model did not overlap with the result from biopsy, the result is still much closer compared to the value assuming same penetration under microdose conditions. This could be explained by the different measurement methods with biopsy and imaging, as well as high variance across different participants^16^.

It’s important to note that the model to estimate prevention efficacy may have a high variance since it was built based on daily blood VRC01 level instead of anorectal tissue. In the future, building an anorectal tissue-based PK/PD model will be needed to verify the relationship.

The study has several limitations: small sample size, the assumption of using the same correlation formula to all the tissues, and assuming the same distribution coefficient in lungs for trastuzumab. These could be addressed with a more comprehensive PBPK model that includes tissue data in animals with concurrent PET studies. Lastly, studies when target is present are planned to evaluate potential impacts of target mediated disposition for the proposed clinical translational studies.

This study found a PET imaging-PBPK approach may be of particular value to inform dosing of bNAbs and other monoclonals where tissue-specific concentration of clinical importance. Further links to clinical prevention efficacy and other clinical outcomes may inform the use of PK/PD targets in drug development.

## Methods

### Sex as a biological variable

As HIV affect patients regardless of sex, men (n=4) and women (n=1) were included in the study cohort.

### Study cohort and PET imaging

Data from participants without HIV from the previous PET imaging study (NCT03729752) was utilized in this study (n=5). Detailed baseline characteristics and study methos are described previously. Briefly, participants were 80% male sex at birth, median age 63 years (range 34-64), and weighing median 189 lbs (range 170-208). The radiolabeled VRC01 (^89^Zr-VRC01) preparation and PET imaging was described previously(21). Briefly, 37 MBq of radiotracer was administered and imaging was performed at 1-2, 4-6, 24, 72, 144 h (n=5 participants with varying timepoints) post tracer injection. Standardized uptake values (SUVs) were determined using 3-dimensional gating on tissue of interest.

### Conversion of radiotracer uptake to concentration

The mean radiolabel uptake signal of the tissue (corrected for label decay), mean standardized uptake value (SUVmean), was plotted against the quantified percent injected drug per gram of blood (%ID/g) at the corresponding time to determine the concentration. The relationship was defined using a linear regression in Excel.

### PET-PBPK model development

We developed a PBPK model adapted from a published model(22) to describe radiolabeled VRC01 pharmacokinetics in blood, liver, gut/anorectal and spleen, based on empirical data and pre-defined physiological parameters (Table 1)(22–26). All models and simulations were constructed and run in NONMEM version 7.5 and/or R version 4.4.0. This includes parameterization for tissue volume (vascular and extravascular space for each organ, V), blood (Q) and lymphatic (L) flow rates, total tissue-based clearance (CL), distribution coefficients, and fractional lymphatic drainage (FR) (Fig. 1). We evaluate proportional, additive, and combined error models to describe residual variability of VRC01 levels in each tissue. The lymphatic capillary reflection coefficient was fixed to 0.2 for all the tissues, and the distribution coefficient between the interstitial space of lungs and blood was fixed to 3.67(22). We evaluated model estimate precision and fit using bootstrap replicates (n=4000) and visual predictive checks (n=1000).

**Table 1.** Estimated parameters of the PBPK model.

| **Variable** | **Point estimate**<br>**(95% bootstrap confidence interval)** |
| --- | --- |
| *Liver-blood distribution coefficient* | 8.06 (5.58 – 8.16) |
| *Gut & Anorectal distribution coefficient* | 1.31 (0.89 – 1.34) |
| *Spleen-blood distribution coefficient* | 1.71 (1.01 – 1.81) |
| *Total clearance, mL/h* | 27.7 (0.28 – 53.5) |
| *Fraction of lymphatic drainage contributing to blood concentration of VRC01 (FR)* | 0.84 (0.72 – 0.97) |
| *Proportional error of blood concentration* | 0.04 (0.01 - 0.08) |
| *Proportional error of gut/anorectal concentration* | 0.06 (0.02 - 0.10) |
| *Proportional error of spleen concentration* | 0.02 (0.01 - 0.03) |
| *Proportional error of liver concentration* | 0.03 (0.01 - 0.05) |

**Table 2.** Literature parameters used in the PBPK model.

| *Variable* | **Human Physiological Model Values** |
| --- | --- |
| *Total blood flow ($Q_b$), mL/h* | 336,000 |
| *Lung blood flow ($Q_{lu}$), mL/h* | 336,000 |
| *Liver blood flow ($Q_h$), mL/h* | 87,000 |
| *Gut & anorectal blood flow ($Q_g$), mL/h* | 66,000 |
| *Spleen blood flow ($Q_s$), mL/h* | 4,620 |
| *Remaining organs blood flow ($Q_o$), mL/h* | 178,380 |
| *Volume of blood ($V_b$), mL* | 5,200 |
| *Lung volume ($V_{lu}$), mL* | 999 |
| *Lung, vascular volume ($V_{luvs}$), mL* | 66.5 |
| *Lung, interstitial space volume ($V_{luis}$), mL* | 199.5 |
| *Liver volume ($V_h$), mL* | 1809 |
| *Liver, vascular volume ($V_{hvs}$), mL* | 350 |
| *Liver, interstitial space volume ($V_{his}$), mL* | 875 |
| *Gut & anorectal volume ($V_g$), mL* | 2,147 |
| *Gut & anorectal, vascular volume ($V_{gvs}$), mL* | 43 |
| *Gut & anorectal, interstitial space volume ($V_{gis}$), mL* | 373.2 |
| *Spleen volume ($V_s$), mL* | 173.4 |
| *Spleen, vascular volume ($V_{svs}$), mL* | 35 |
| *Spleen, interstitial space volume ($V_{sis}$), mL* | 70 |
| *Remaining organs volume ($V_o$), mL* | 59,652 |
| *Remaining organs, intersistial space volume ($V_{ois}$), mL* | 12,524.3 |
| *Lung lymphatic flow ($L_{lu}$), mL/h* | 38.5 |
| *Liver lymphatic flow ($L_h$), mL/h* | 52.5 |
| *Gut & anorectal lymphatic flow ($L_g$), mL/h* | 49.8 |
| *Spleen lymphatic flow ($L_s$), mL/h* | 20.3 |
| *Remaining organs lymphatic flow ($L_o$), mL/h* | 475.24 |

One of the parameters – distribution coefficient is defined as the ratio of extravascular space concentration of VRC01 and blood. As antibodies are large molecules, the intracellular space concentration was considered to be negligible(27).

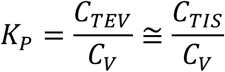

Kp: Distribution coefficient
C_TEV_: Extravascular (interstitial and intracellular) space of the tissue
C_TIS_: Interstitial space of the tissue

The tissue distribution rate (except other tissue) consisted of two parts – blood flow and lymphatic flow, where the equations were defined as below:

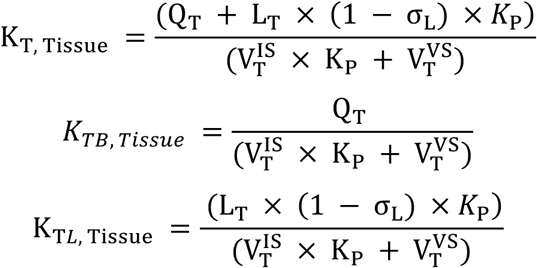

T: corresponding tissue
Q_T_: blood flow to the tissue
L_T_: lymphatic flow to the tissue, K

### Simulation of clinical trial for clinical dose administration

The developed PBPK model was used to simulate with two cohorts consisting of 1000 70-kg adult virtual participants each, administered with 10 doses of 10mg/kg and 30mg/kg, respectively, every 8 weeks to match the AMP trials(18). The mean VRC01 concentration and 90% prediction interval was for each dosing cohort was plotted.

### Statistics

The predicted daily concentration of VRC01 was derived using the population pharmacokinetic (popPK) model established by Gilbert and colleagues(19). In conjunction with this, the non-human primates model developed by Pegu and colleagues was applied to estimate the prevention efficacy by assessing the reduction in risk per day(28, 29).

We then modeled the prevention efficacy, using a log-logistic function with the predicted anorectal concentration divided by AMP trial observed IC_80_ values. For the PET-based predicted anorectal concentrations, we investigated two approaches: First using ratios based on the mean, minimum, and maximum observed PET SUVmean signal in tissue:blood, second, using PBPK model predicted mean anorectal:blood levels and the predicted 90% prediction interval. Using the aforementioned model of prevention efficacy versus plasma VRC01 level:IC_80_, we used the derived ratios to predict corresponding prevention efficacy and the tissue VRC01:IC_80_ ratio needed for 90% prevention efficacy. All analysis were performed using R version 4.4.0.

### Study approval

The US FDA approved the use of ^89^Zr-VRC01 under IND 139652. The UCSF Committee on Human Research Institutional Review Board and the UCSF Radiation Safety Committee approved the study. Written informed consent was obtained from each participant prior to study entry. ClinicalTrial.gov: NCT03729752.

## Data availability

Primary PET data was previously published by Beckford-Vera et al.(21). The radiotracer and corresponding VRC01 concentration in blood is attached in Supplemental Document 1. The AMP trial data were utilized from the pharmacokinetics data previously published by Gilbert et al.(19) and Huang et al.(32) on the data shared in ATLAS Science Portal (https://atlas.scharp.org/project/home/begin.view). The NONMEM code used for the PBPK model is attached in Supplemental Document 2.

## Acknowledgements

This work is supported by the National Institutes of Health R01AI52932 (to T.J.H., H.F.V.), K23AI162249 (to A.N.D.), K24AI174971 (to T.J.H.), and P30AI027763 (CFAR Resource Allocation Program to A.N.D.). The content is solely the responsibility of the authors and does not necessarily represent the official views of the National Institutes of Health.

## Author Contributions

D.H.C., L.K.E., and A.N.D. conceived and designed the study, analyzed data and wrote the manuscript. D.R.B., R.R.F., Y.S., H.F.V., and T.J.H performed study for PET imaging, generated output data, advised on translational analysis and edited the manuscript.

